# From Binge Drinking to Future Alcohol Severity: The Role of Emotion Regulation and Emerging Psychopathology

**DOI:** 10.1101/2025.03.04.25323343

**Authors:** Carina Carbia, María Soledad Rodríguez, Samuel Suárez-Suárez, Sonia Doallo, Fernando Cadaveira, Montserrat Corral

## Abstract

Binge drinking (BD) during emerging adulthood increases the risk of developing alcohol use disorders, yet not all individuals follow this trajectory. Deficits in emotion regulation has been identified as a risk factor for psychopathology, but its specific role in shaping long-term alcohol severity among young binge drinkers remains insufficiently understood. This study investigates the role of emotion regulation difficulties as a mediator in the relationship between binge BD and future alcohol severity in young people and the moderator role of emerging psychopathological symptoms. We followed a cohort of 192 university students (53% female) over two years, from ages 18 to 20 We measured alcohol consumption and emotion regulation (Difficulties in Emotion Regulation Scale), as well as psychopathological symptoms (Brief Symptom Inventory). Mediation and moderated mediation models were conducted with PROCESS. Results show that BD predicts future alcohol severity, with emotion regulation difficulties, specifically challenges in goal-directed behaviour, partially mediating this relationship. Psychopathological symptoms moderated the effects, with difficulties in emotion regulation being a significant predictor of future alcohol severity only in individuals with emerging psychopathological symptoms. Additionally, the association between BD and future alcohol severity was amplified in those with emerging psychopathological symptoms. These findings offer new insights into the risk factors underlying the escalation of problematic alcohol use and the interplay between BD, emotion regulation, and psychopathology. They also stress the importance of early interventions focused on enhancing emotion regulation abilities in young binge drinkers, especially those displaying early psychopathological symptoms.

## 1. INTRODUCTION

Alcohol consumption is the leading risk factor for premature mortality among individuals aged 15 to 49 (World Health Organization [WHO], 2018). The most prevalent pattern of alcohol use among young adults is binge drinking (BD), typically defined as consuming a considerable quantity of alcohol in a short period that results in a blood alcohol concentration of at least 0.8 g/L (National Institute on Alcohol Abuse and Alcoholism, 2004). BD peaks in late adolescence, particularly in the early 20s, coinciding with college years (Merrill & Carey, 2016). While alcohol use generally declines during emerging adulthood, a significant subset of individuals continues to binge drink into adulthood (Moure-Rodríguez et al., 2018). These persistent binge drinkers (BDs) face a heightened risk of developing alcohol use disorders (AUDs) (Patrick et al., 2021). Understanding the factors driving the progression from BD to problematic alcohol use during the critical transition to early adulthood is essential for better identification of at-risk subgroups. Since substance use and comorbid psychopathology peak during adolescence, it is crucial to focus on transdiagnostic risk factors underlying the frequent co-occurrence of alcohol misuse and mood disorders to develop effective prevention strategies that target multiple disorders (Compas et al., 2017; González-Roz et al., 2024; Shadur & Lejuez, 2015).

In this sense, one risk factor that has gained attention over the last years due to its potential transdiagnostic value is emotion regulation (Klein et al., 2022; Lincoln et al.; 2022; McLaughlin et al, 2015; Schäfer et al., 2017). Emotion regulation refers to individuals’ efforts to influence which emotions they experience and express, effectively adapt to different situations, and achieve desired emotional states (Gross, 1998). Deficits in emotion regulation have been previously demonstrated in patients with AUD, being more consistently associated with alcohol use problems than frequency of alcohol use (Brown & Melas, 2024). A recent meta-analysis (Stellern et al., 2023) concluded that individuals with AUD exhibit difficulties in different emotion regulation dimensions, principally abilities related to impulsive behaviour, lack of adaptive strategies and difficulties engaging in goal-directed behaviour. These difficulties are linked to higher relapse rates (Fox et al., 2008; Shadur & Lejuez, 2015) and appear to contribute to higher alcohol craving (Khosravani et al., 2017; Petit et al., 2015; You et al., 2023).

Thus, while poor emotion regulation increases alcohol misuse, chronic alcohol use further impairs an individual’s ability to regulate emotions (Brown & Melas, 2024). According to the negative reinforcement model of addiction, individuals may use substances as a maladaptive coping mechanism to alleviate negative emotions, thereby reinforcing the cycle of substance use and emotional dysregulation (Koob, 2013; Wise & Koob, 2014). Consequently, individuals with AUD and comorbid psychopathology exhibit even more severe deficits in emotion regulations, with greater reliance on substance use as an ineffective emotion regulation strategy (Berking et al., 2011; Shadur & Lejuez, 2015).

Given that adolescence is a critical period for the onset of psychiatric disorders (Compas et al., 2017; McLaughlin et al., 2011; Rapee et al., 2019), difficulties in emotion regulation during this stage may increase vulnerability to substance use. This developmental period is characterized by heightened social changes and increased stressors. Moreover, significant neuromaturational changes are taking place, particularly in brain regions underlying self-control and emotion regulation, such as the prefrontal cortex and limbic system (Ahmed et al., 2015) and continue to develop through adolescence into emerging adulthood (Andrews et al., 2021; Fombouchet et al., 2023). Thus, adolescents who engage in BD and lack adaptive emotion regulation, might be at heightened risk for future alcohol severity and alcohol-related problems.

Despite this, limited research has explored the role of emotion regulation among young BDs and its impact on future alcohol severity. Few longitudinal studies have examined how emotion regulation and substance use interact over time. For example, Hessler and Katz (2010) found that adolescents with poor emotion regulation were more likely to engage in substance use seven years later. Even after controlling for baseline substance use, emotion dysregulation predicted future emergence of mood disorders and greater substance use, strengthening claims of emotion regulation as a key transdiagnostic risk factor (Klein et al., 2022; Kliewer et al., 2016).

Building on existing evidence, this longitudinal study examines whether difficulties in emotion regulation mediate the relationship between BD and trajectories of alcohol severity in university students over a two-year period. Specifically, we hypothesize that BD will predict greater alcohol severity over time, with this relationship being partially explained by emotion regulation difficulties (mediation hypothesis). Furthermore, we propose that psychopathological symptoms will serve as a moderator, influencing the strength of these associations (moderated mediation hypothesis). Specifically, we expect that young BDs with higher levels of psychopathological symptoms will exhibit a stronger relationship between emotion regulation difficulties and future alcohol severity, suggesting that difficulties in managing emotions may be particularly detrimental for those individuals who experience greater psychopathological symptoms. Additionally, we hypothesize that psychopathological symptoms will moderate the direct effect of BD on alcohol severity, such that the association will be stronger for those with greater psychopathological symptoms. By examining these interconnections, this study aims to provide a more nuanced understanding of the mechanisms underlying the risk of escalating alcohol use severity and highlight the potential role of emotion regulation and comorbid psychopathology in shaping these trajectories.

## 2. MATERIALS AND METHODS

### 2.1. Sample and Procedures

The sample consisted of 192 participants (102 females) who were recruited as part of a longitudinal study conducted within the scope of a larger research project investigating risk factors and outcomes associated with BD in university students. At baseline, the sample was composed by 267 participants and 192 at follow-up (two years later). For sample selection, a total of 2998 first-year students from the Universidade de Santiago de Compostela (USC, Spain) voluntarily completed a questionnaire assessing sociodemographic information and alcohol and other substance consumption, including the adapted versions of the Alcohol Use Disorders Identification Test (AUDIT; Babor et al., 2001; Bohn et al., 1995; Varela et al., 2005), Timeline Follow-Back (TLFB; Sobell & Sobell, 1992), and the Cannabis Abuse Screening Test (CAST; Cuenca-Royo et al., 2012; Legleye et al., 2007). Those interested in participating provided a telephone number/email and were scheduled for an initial visit. At this initial visit, the eligibility for study participation was based on the following exclusion criteria: potential alcohol-related problems (>20 AUDIT); psychopathological symptoms (≥ 90th percentile GSI score, Symptom Check-list-90 SCL-90, Derogatis & Savitz,1999); neurological or systemic conditions that significantly impacted neurocognitive function; regular use of psychoactive medications; consumption of illicit substances in the las 6 months, except occasional cannabis (CAST <9); and family history of alcoholism or a diagnosed major psychopathological disorder (defined as at least two first-degree relatives or three or more first- or second-degree relatives with psychiatric history). Participants reported their alcohol consumption at two different times: at baseline and follow-up, when they were between 18/19 years old and between 20/21 years old. Emotion regulation was assessed at follow-up, with the Spanish validation of the Difficulty in Emotion Regulation Scale (DERS; Gratz & Roemer, 2004; Hervás & Jódar, 2008). Additionally, at follow-up, psychopathological symptoms were measured using the Brief Symptom Inventory (BSI-18; Derogatis, 2001; Derogatis, 2013). All participants gave written consent and received monetary compensation for their participation. The study received approval from the USĆs Bioethics Committee.

### 2.2. Materials

#### Difficulty in Emotion Regulation Scale

(DERS; Gratz & Roemer, 2004): This self-report questionnaire assesses difficulties with emotion regulation. The DERS assesses six different aspects of emotional regulation, including non-acceptance of emotional responses (Non-Acceptance), lack of emotional awareness (Awareness), impulse control difficulties (Impulse), difficulty engaging in goal-directed behaviour (Goals), lack of emotional clarity (Clarity) and limited access to emotional regulation strategies (Strategies). Non-acceptance of emotional responses examines the extent to which individuals experience additional negative emotion as a result their evaluation of current emotional states (e.g. ‘‘When I’m upset, I become irritated with myself for feeling that way’’). Lack of emotional awareness indexes the extent to which individuals attend to and acknowledge their emotions (‘‘I am attentive to my feelings’’). Lack of emotional clarity indexes difficulties in identifying which emotions a person is experiencing or confusion about emotional experiences (e.g., ‘‘I have difficulty making sense out of my feelings’’). Two domains focus on the extent to which emotions affect goal-directed behaviour. Impulse control difficulties index the extent to which negative emotion increases the likelihood of rash/impulsive action at the expense of future goals (e.g. ‘‘When I’m upset, I become out of control’’). Difficulties engaging in goal-directed behaviour indexes the extent to which negative emotions interrupt an individual’s ability to maintain focus on specific tasks (e.g., ‘‘When I’m upset, I have difficulty getting work done’’). Finally, limited access to emotion regulation strategies, focuses on an individual’s subjective belief in their ability to use emotion regulation when needed. The total DERS score is calculated from the sum of all six subscales (range of 36 to 180), with higher scores indicating higher difficulties. The DERS has shown good internal consistency (Cronbach’s α > 0.70); similar values were obtained in our study (0.93 Cronbach’s α) with adequate predictive and construct validity (Hervás & Jódar, 2008).

#### Alcohol Use Disorders Identification Test

(AUDIT; Babor et al., 2001): The AUDIT is a screening tool developed by the WHO to identify problem drinkers, which includes questions on alcohol use, dependence symptoms, and alcohol-related problems over the past 12 months. In this study, the Galician-validated version was used. Scores range from 0 to 40 (high risk of alcohol dependence). The Cronbach’s alpha in this sample was 0.812. The total AUDIT score at follow-up was used as an index of alcohol severity in the models.

#### Alcohol Timeline Follow-Back calendar

(TLFB; Sobell & Sobell, 1992): Participants retrospectively completed a calendar for the past 180 days, recording the number of standard alcohol units consumed each day. This measure allowed us to compute the number of BD days (over the past 180 days), defined as the consumption of ≥ 4 (female) or ≥ 6 (male) Spanish standard drinks (10g of alcohol) on one single occasion. Number of BD episodes in the past 180 days was used in the statistical models as the main predictor.

#### Brief Symptom Inventory 18

(BSI-18; Derogatis, 2001; Derogatis, 2013): The BSI is a self report questionnaire with 18 items answered on a Likert scale. Considered a shortened version of the SCL-90, it assesses psychopathological symptoms in both clinical and general populations. It measures factors like Somatization, Depression, and Anxiety and a Global Severity Index, which was used as an indicator of emerging psychopathological symptoms at follow-up.

### 2.3. Statistical Analysis

Descriptive statistics, including demographic data, alcohol consumption, BSI-18 and DERS scores were calculated. T tests were used to assess sex-related differences and to assess differences between groups with high and low psychopathological symptoms after a moderation effect was proven. Correlations between DERS subscales and drinking variables were computed. Mediation and moderated mediation analyses were conducted with the PROCESS macro (Hayes, 2012; Hayes, 2017), using the IBM Statistical Package SPSS Version 28.0.

The assumptions for the Mediation Model were as follows: binge drinking influences future alcohol severity both directly and indirectly through emotion regulation difficulties. Specifically, we propose: (I) BD will directly predict alcohol severity and (II) the effect of BD on future alcohol severity will be mediated by emotion regulation. The assumptions for the Moderated Mediation in which psychopathological symptoms were proposed as moderator are as follows: (I) The strength of the association between emotion regulation difficulties and future alcohol severity will be moderated by psychopathological symptoms, such that this relationship will be stronger for individuals with higher levels of psychopathological symptoms; (II) The direct effect of BD on future alcohol severity will also be moderated by psychopathological symptoms, such that the association between BD and alcohol severity will be stronger among individuals with higher levels of psychopathological symptoms. Sample categorization in high and low psychopathological symptoms was performed using the median split (BSI score at follow-up, 50th percentile).

## 3. RESULTS

### 3.1. Descriptive statistics and correlations

Descriptive statistics for the full sample are shown in Table 1. When comparing low and high psychopathology groups, individuals with higher psychopathological symptoms exhibited more alcohol use severity at follow-up and more pronounced difficulties in the total DERS score, and all dimensions except Awareness. However, no significant differences between these two groups were observed for the number of BD episodes at baseline. Correlations between study variables can be found in Table 2. The number of BD episodes positively correlated with severity of alcohol use as measured by the AUDIT total at follow-up (r = .638), as well as with the total DERS score (r= .171). Among the DERS dimensions, the Goals dimension showed the strongest correlation with the number of BD days (r = .191) and AUDIT total (r = .300).

**Table 1.** Descriptive characteristics of the total sample and psychopathology groups (Low vs. High) (Mean ± SD)

|  | Total | Low-P | High-P |
| --- | --- | --- | --- |
| N (females) | 192 (102) | 100 (46) | 92 (56) |
| BD days<sup>a</sup> | 15.39 (± 15.91) | 14.26 (± 16.11) | 16.63 (± 15.69) |
| AUDIT<sup>b</sup> | 6.38 (± 5.18) | 5.08 (± 3.86) | 7.79 (± 6.01)*** |
| BSI-18 | 0.49 (± 0.37) | 0.22 (± 0.14) | 0.79 (± 0.31)*** |
| DERS Total | 77.15 (± 19.22) | 68.02 (± 13.95) | 87.08 (± 19.32)*** |
| Non-Acceptance | 13.46 (± 5.52) | 11.41 (± 4.11) | 15.68 (± 6)*** |
| Awareness | 15.42 (± 4.08) | 14.9 (± 3.73) | 15.98 (± 4.38) |
| Impulse | 10.75 (± 4.54) | 9.35 (± 3.66) | 12.27 (± 4.93)*** |
| Goals | 14.29 (± 4.48) | 12.78 (± 4.35) | 15.93 (± 4.04)*** |
| Clarity | 10.05 (± 3.34) | 8.56 (± 2.45) | 11.67 (± 3.46)*** |
| Strategies | 13.23 (± 5.12) | 11.02 (± 3.20) | 15.63 (± 5.71)*** |

<sup>a</sup> Measured at baseline, relative to the six prior months using the alcohol TLFB. <sup>b</sup> Measured at follow-up. Low-P= Low Psychopathological symptoms. High-P = High Psychopathological Symptoms. \*\*\* $p < 0.001$ , as per T-test between groups of Low and High psychopathology at follow-up, based on median split (percentile 50).

**Table 2.**
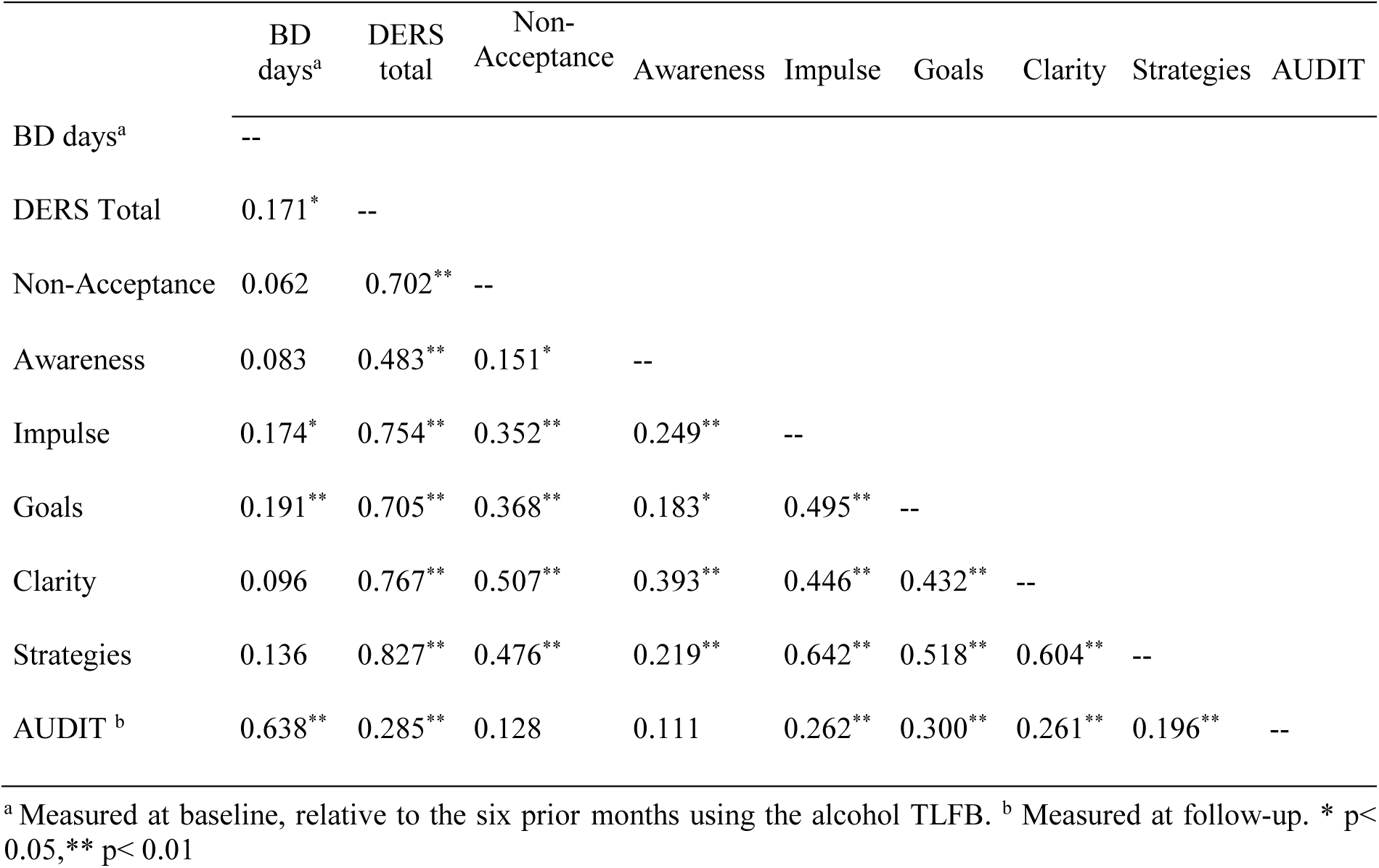
Correlation values between DERS scores and alcohol consumption variables at baseline and at follow-up.

|  | BD<br>days<sup>a</sup> | DERS<br>total | Non-Acceptance | Awareness | Impulse | Goals | Clarity | Strategies | AUDIT |
| --- | --- | --- | --- | --- | --- | --- | --- | --- | --- |
| BD days<sup>a</sup> | -- |  |  |  |  |  |  |  |  |
| DERS Total | 0.171<sup>*</sup> | -- |  |  |  |  |  |  |  |
| Non-Acceptance | 0.062 | 0.702<sup>**</sup> | -- |  |  |  |  |  |  |
| Awareness | 0.083 | 0.483<sup>**</sup> | 0.151<sup>*</sup> | -- |  |  |  |  |  |
| Impulse | 0.174<sup>*</sup> | 0.754<sup>**</sup> | 0.352<sup>**</sup> | 0.249<sup>**</sup> | -- |  |  |  |  |
| Goals | 0.191<sup>**</sup> | 0.705<sup>**</sup> | 0.368<sup>**</sup> | 0.183<sup>*</sup> | 0.495<sup>**</sup> | -- |  |  |  |
| Clarity | 0.096 | 0.767<sup>**</sup> | 0.507<sup>**</sup> | 0.393<sup>**</sup> | 0.446<sup>**</sup> | 0.432<sup>**</sup> | -- |  |  |
| Strategies | 0.136 | 0.827<sup>**</sup> | 0.476<sup>**</sup> | 0.219<sup>**</sup> | 0.642<sup>**</sup> | 0.518<sup>**</sup> | 0.604<sup>**</sup> | -- |  |
| AUDIT<sup>b</sup> | 0.638<sup>**</sup> | 0.285<sup>**</sup> | 0.128 | 0.111 | 0.262<sup>**</sup> | 0.300<sup>**</sup> | 0.261<sup>**</sup> | 0.196<sup>**</sup> | -- |

<sup>a</sup> Measured at baseline, relative to the six prior months using the alcohol TLFB. <sup>b</sup> Measured at follow-up. \* p< 0.05, \*\* p< 0.01

### 3.2. Mediation analysis

The results indicate that while the number of BD episodes is the strongest predictor for later alcohol use severity, there was a significant indirect effect mediated by emotion regulation. The model explains 44% of the variance in predicting future alcohol severity two years later. This association was mediated by the Goals dimension, that is, difficulties engaging in goal-directed behaviour. Table 3 contains the coefficients of all regression-based models. A conceptual representation can be found in Figure 1.

**Figure 1.**
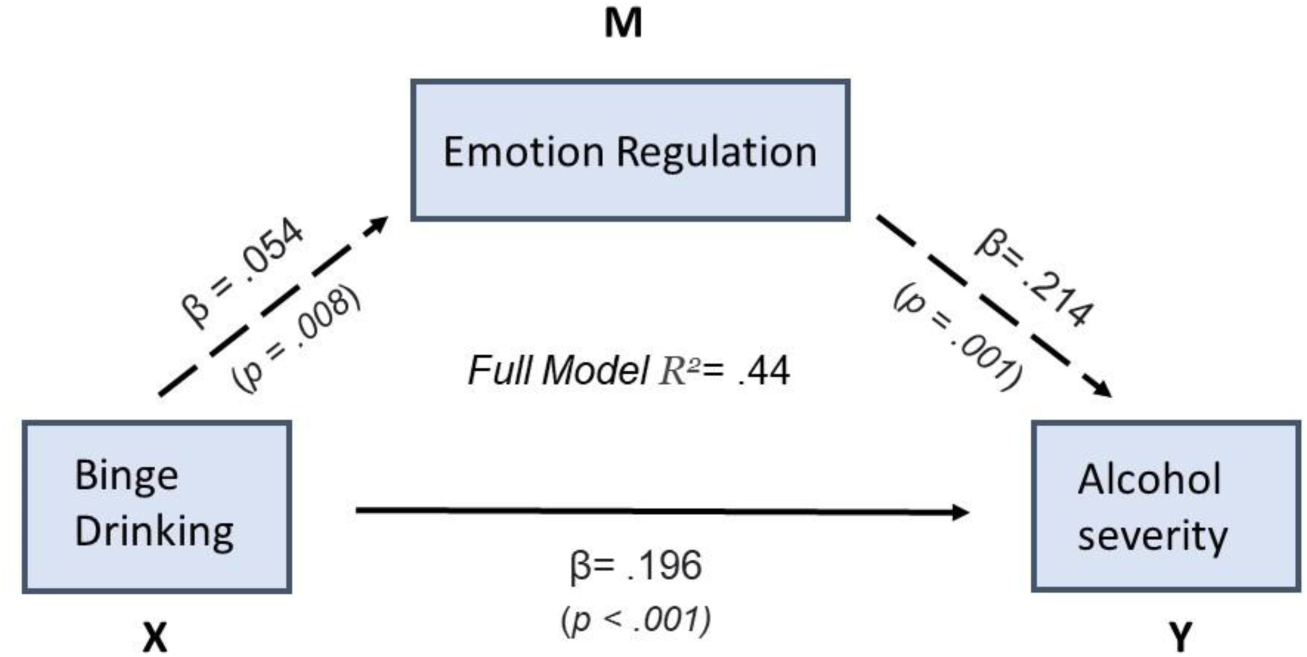
Statistical parameters for the Mediation Model

**Table 3.** Regression Coefficients for Mediation and Moderated Mediation Models.

| Pathway | Predictor | B | SE | t | p | 95% CI<br>(LL, UL) |
| --- | --- | --- | --- | --- | --- | --- |
| **Mediation Model** |  |  |  |  |  |  |
| **M → Y** | Emotion Regulation → Alcohol Severity | 0.214 | 0.064 | 3.338 | .001 | [0.088, 0.340] |
| **X → M** | Binge Drinking → Emotion Regulation | 0.054 | 0.020 | 2.683 | .008 | [0.014, 0.093] |
| **X → Y (Total Effect)** | Binge Drinking → Alcohol Severity | 0.208 | 0.018 | 11.422 | <.001 | [0.172, 0.244] |
| **X → Y (Direct Effect)** | Binge Drinking → Alcohol Severity | 0.196 | 0.018 | 10.869 | <.001 | [0.161, 0.232] |
| **Indirect Effect (X → M → Y)** | Binge Drinking → Emotion Regulation → Alcohol Severity | 0.012 | 0.005 | — | — | [0.002, 0.024] |
| **Moderated Mediation Model** |  |  |  |  |  |  |
| **X → Y (Direct Effect, Moderated by BSI)** | Binge Drinking → Alcohol Severity | 0.150 | 0.023 | 6.476 | <.001 | [0.105, 0.196] |
| **X × W Interaction (Moderation of Direct Effect)** | Binge Drinking × Psychopathology | 0.095 | 0.034 | 2.768 | .006 | [0.027, 0.162] |
| **M × W Interaction (Moderation of Indirect Effect)** | Emotion Regulation × Psychopathology | 0.301 | 0.130 | 2.309 | .022 | [0.044, 0.558] |
| **Conditional Effect of X on Y at Low BSI** | Binge Drinking → Alcohol Severity (Low BSI) | 0.150 | 0.023 | 6.476 | <.001 | [0.105, 0.196] |
| **Conditional Effect of X on Y at High BSI** | Binge Drinking → Alcohol Severity (High BSI) | 0.245 | 0.025 | 9.732 | <.001 | [0.195, 0.295] |
| **Conditional Effect of M on Y at Low BSI** | Emotion Regulation → Alcohol Severity (Low BSI) | 0.006 | 0.086 | 0.069 | .945 | [-0.164, 0.175] |
| **Conditional Effect of M on Y at High BSI** | Emotion Regulation → Alcohol Severity (High BSI) | 0.307 | 0.098 | 3.134 | .002 | [0.114, 0.500] |

Notes: B = unstandardized coefficient, SE = standard error, t = t-value, p = significance level, 95% CI = confidence interval (lower and upper bounds). Low BSI= Low psychopathological symptoms based on median split (percentile 50). High BSI= Low psychopathological symptoms based on median split (percentile 50).

BD significantly predicted emotion regulation difficulties (B = .054, p = .008), indicating that higher number of BD episodes was associated with greater difficulties in emotion regulation, in particular difficulties in goal-directed behaviour. The overall model was significant (F_2,189_ = 74.283, p < .001, R² = .440) and explained 44.0% of the variance. Both BD (B = .196, p < .001) and emotion regulation difficulties in Goals (B = .214, p = .001) were significant predictors of alcohol severity, with a significant indirect effect of BD on alcohol severity through the Goals dimension (B = .012, 95% CI = .002 - .024), supporting mediation. These findings suggest that emotion regulation difficulties in goal-directed behaviour, partially explain the association between BD and alcohol severity, while BD still retains a significant direct influence on future alcohol severity.

### 3.3 Moderated Mediation analysis

This model examined whether psychopathological symptoms at follow-up moderated both the indirect effect and the direct effect of BD on future alcohol severity. The overall model was significant (F_5,186_ = 38.484, p < .001), explaining 51% of the variance. Table 3 contains the coefficients of all regression-based models.

BD remained a significant predictor (B = .150, p < .001). The interaction between BD and psychopathological symptoms was significant (B = .095, p = .006), indicating that psychopathological symptoms moderate the direct effect of BD on alcohol severity. The interaction between emotion regulation difficulties and psychopathological symptoms was also significant (B = .301, p = .022).

At low levels of psychopathological symptoms, the effect of emotion regulation on alcohol severity was non-significant. However, at high levels of psychopathological symptoms the effect of emotion regulation on alcohol severity was significant (B = .307, p = .002), indicating that difficulties in emotion regulation only significantly predicted alcohol severity for those with high psychopathology. At low levels of psychopathological symptoms, the effect of BD on alcohol severity was lower than at high levels (B = .150, p < .001). At high levels of psychopathological symptoms, this effect was stronger (B = .245, p < .001), suggesting that BD had a stronger association with future alcohol severity among individuals that develop psychopathological symptoms. Figure 2 depicts a conceptual representation for the overall model and psychopathology groups.

**Figure 2.**
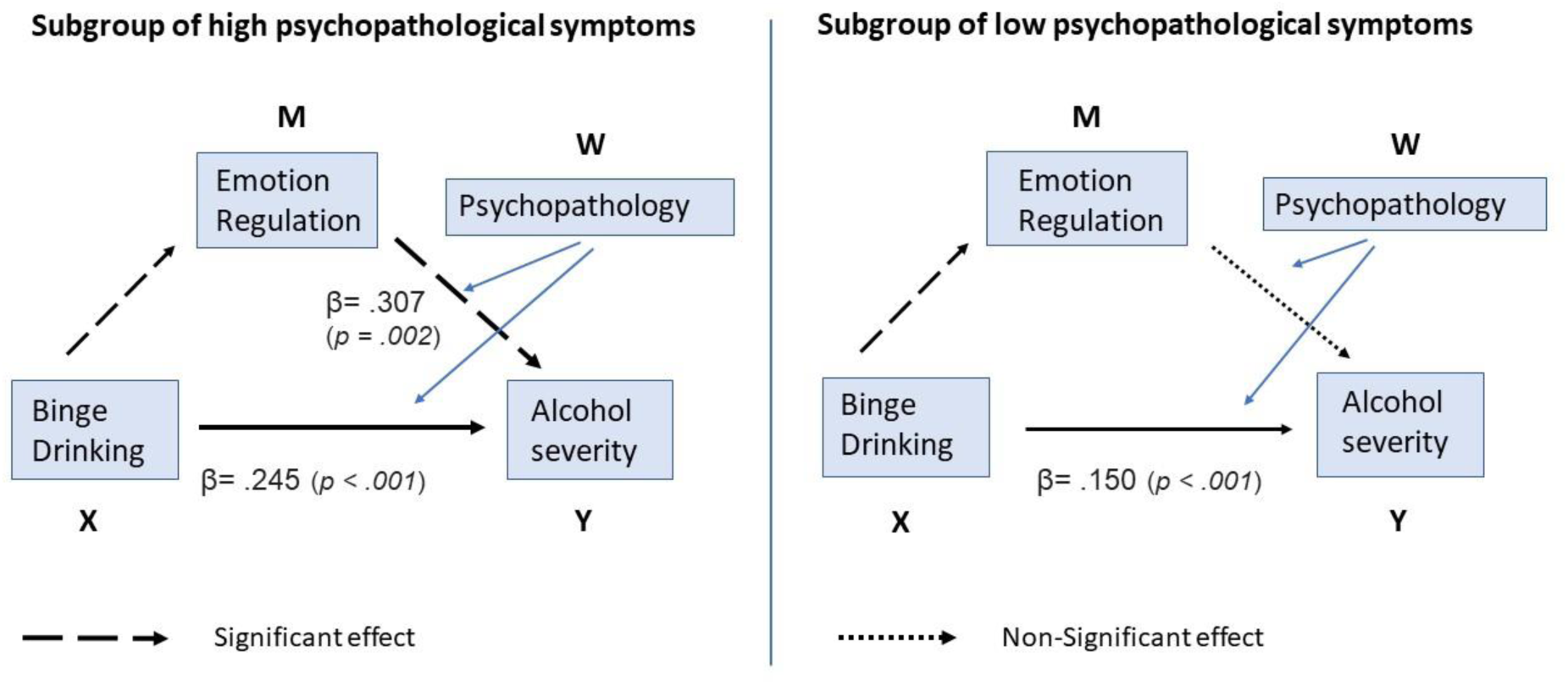
Statistical parameters for the Moderated Mediation model

## 4. DISCUSSION

While emotion dysregulation is known to contribute to alcohol dependence, few studies have explored its role in young non-clinical samples, such as university students who regularly binge drink. This study examines how emotion regulation mediates the link between BD and future alcohol severity over two years in young people without prior alcohol use disorders or psychopathology. First, our findings show that a higher number of BD episodes was associated with greater difficulties in emotion regulation. As hypothesized, while BD remained a significant predictor of future alcohol severity, emotion regulation difficulties partially mediated this relationship. Specifically, difficulties engaging in goal-directed behaviour under distress—such as struggling to concentrate and complete tasks when experiencing negative emotions—emerged as the key dimension associated with this mediation effect. Although our sample excluded individuals with psychopathological disorders at baseline, emerging psychopathological symptoms at follow-up significantly moderated these associations. Thus, among individuals with high psychopathological symptoms, BD was a stronger predictor of alcohol severity than at low levels of symptomatology. Additionally, emotion regulation difficulties predicted alcohol severity only in those with high psychopathological symptoms.

These results align with previous research demonstrating that emotion dysregulation is a critical risk factor for substance use disorders (SUDs) (Cavicchioli et al., 2023; Fox et al., 2008; Weiss et al., 2022). Individuals with SUDs exhibit poorer emotion regulation across all DERS subscales, with Strategies and Impulse subscales showing the largest deficits (Stellern et al., 2023). In view of this difficulties, substance use may serve as a maladaptive strategy to cope with negative emotions (Stellern et al., 2023). In AUD patients, pre-treatment emotion regulation skills predicted alcohol use during treatment, while post-treatment skills predicted use at follow-up (Berking et al., 2011) and they are also linked to increased craving and relapse vulnerability, even when controlling for other factors (Fox et al., 2008; Khosravani et al., 2017; Petit et al., 2015; You et al., 2023). A recent meta-analysis showed that difficulties in reducing the intensity and duration of negative emotions are associated with alcohol-related problems and severity of the disorder (González-Roz et al., 2024).

In non-clinical samples of university students, high levels of emotion regulation difficulties appear to be linked to a greater risk of alcohol-related problems (Benzerouk et al., 2022; Kim & Kwon, 2020). In particular, the DERS Impulse and Goals dimensions were associated with drinking speed and BD episodes, respectively (Benzerouk et al., 2022). These dimensions reflect difficulties in disengaging from preoccupying affective states, which is linked to cognitive control deficits (Gross, 2015; McRae & Gross, 2020), a brain network still undergoing significant refinement until adulthood (Malagoli et al., 2022; Poon et al., 2016). Similarly, Dvorak and colleagues (2014), using the DERS scale, demonstrated that impulse control difficulties were positively related to the number of drinks consumed per week, while difficulty engaging in goal-directed behaviour was associated with experiencing greater alcohol-related consequences. Among young heavy drinkers, difficulties regulating both positive and negative emotions appear to be more relevant as risk factors for alcohol problems than actual alcohol consumption (Paulus et al., 2021). In this sense, limited access to emotion regulation strategies significantly predicted alcohol-related problems via depression and anxiety coping motives but did not predict alcohol consumption (Simons et al., 2017). Overall, research suggests that goal-directed self-regulation deficits are associated with alcohol-related consequences rather than alcohol use itself among college students (Teeters et al., 2023; Brown & Melas, 2024). Poor emotion regulation likely not only predicts greater alcohol use severity but is also worsened by continued drinking, creating a self-perpetuating cycle (Lannoy et al., 2021; Martini et al., 2025).

In our study, emerging psychopathological symptoms moderated the relationship between BD, emotion regulation difficulties, and future alcohol severity, explaining 51% of the variance at follow-up. This suggests that emotion regulation difficulties and psychopathology jointly heighten addiction risk, even in those without prior disorders. These findings align with previous research showing that individuals with both substance use disorders and psychopathology experience greater emotion regulation deficits and more severe substance use (Berkin et al., 2011, Weiss et al., 2022). In a sample of 1,262 adolescents, greater emotion regulation ability at baseline predicted lower psychopathology and substance dependence seven years later (Klein et al., 2022), strengthening evidence of emotion regulation as a transdiagnostic risk factor (Shadur & Lejuez, 2015).

While this study provides valuable insights, it has several limitations. Although the longitudinal design strengthens temporal validity, causal interpretations should be made cautiously. Additionally, self-reported emotion regulation difficulties may not fully capture the complexities of real-world regulation processes. Future research should incorporate ecological momentary assessments and examine emotion regulation flexibility (Chen et al., 2024, English & Eldesouky, 2020) for a more nuanced understanding of these mechanisms and the trajectory of alcohol use.

In conclusion, our study highlights that difficulties in goal-directed behaviour under negative emotions partially mediate the link between BD and future alcohol severity, with emerging psychopathological symptoms amplifying the effects. These findings highlight the crucial role of BD—the most common pattern of alcohol misuse in adolescents—and emotion regulation in the escalation of alcohol use during emerging adulthood. Furthermore, they underscore the need for early interventions to strengthen emotion regulation skills in young BDs, especially those with early signs of psychopathological symptoms.

## Data Availability

All data produced in the present study are available upon reasonable request to the authors

## Acknowledgements

This work was supported by Grant PID2020-113487RB-I00 funded by MCIU/AEI/ 10.13039/501100011033; Grant PNSD 2015/034 funded by PNSD; Grant PSI2015-70525-P co-funded by MEIC and ERDF. Carina Carbia received funding from the Fonds de la Recherche Scientifique (FNRS) as a Chargé de Recherches (CR).

